# A Single Center Observational Study on 25-hydroxyvitamin D levels in Meniscus Injury Patients

**DOI:** 10.1101/2024.08.13.24311955

**Authors:** Shuaishuai Hu, Duzheng Zhang, Ruijun Cong

**Author notes:** **Corresponding authors:** Shuaishuai Hu,; Ruijun Cong. These authors contributed equally to this work (Shuaishuai Hu, Ruijun Cong). These authors contributed equally to this work (Shuaishuai Hu, Duzheng Zhang).

## Abstract

There is limited research on the association between 25-hydroxyvitamin D levels and meniscus injury. This study investigated serum 25-hydroxyvitamin D (25-OHD) in meniscus injury patients and its association with other bioinorganic chemistry of micronutrients, and immune cells numbers from January 2023 to December 2023. A total of 198 participants were recruited between the age of 18 and 89 years. Participants with renal or liver failure, or any other chronic conditions, who were taking medications that might affect the metabolism of vitamin D, were not included in the study. In this study, we categorized serum 25(OH)D concentrations as follows: ≤30ngm/dl was categorized as insufficient, and >30ngm/dl was deemed sufficient. Among the 198 participants, 82% (n=162) were serum 25(OH)D deficient, while only 18% (n=36) participants were serum 25(OH)D sufficient. No significant difference observed in vitamin D deficiency among difference age, BMI, sex, blood pressure, inflammatory cell numbers, and other vitamins compared to the vitamin D sufficient group. Moreover, the serum 25(OH)D concentrations were negatively related to the severity of meniscus injury based on MRI examination. In conclusion, severe vitamin D deficiency is more common in patients with meniscus injury and may play a significant role in their prognosis.

## 1. Introduction

Meniscus injury is one of the most common knee injuries, which occur at an estimated rate of about 60 per 100,000 individuals [1]. The menisci are two crescent-shaped cartilage structures positioned between the femur and tibia in the knee joint. They play a critical role in maintain knee stability by absorbing shock, distributing load, and facilitating smooth joint movement [2-4]. Due to the high incidence of meniscus injuries, understanding the nature and the impact is crucial for reducing the incidence of meniscus injuries.

Recent studies have emphasized the significance of nutritional factors in musculoskeletal and joint health, particularly focus on vitamin D [5-7]. Vitamin D is essential for calcium absorption and bone mineralization [8-9]. However, its impact goes beyond bone health, affecting muscle function and overall joint health [10-12]. Vitamin D deficiency is widespread globally and has been associated with several musculoskeletal disorders, such as osteoporosis and muscle weakness [13-15], potentially increasing the risk and severity of meniscus injuries. However, there is no directly evidence to document the association between vitamin D and meniscus injuries, there is a need for studies to understand the interplay between vitamin D deficiency and meniscus health to develop comprehensive prevention and treatment strategies.

This study aimed to examine the levels of 25-hydroxy vitamin D in patients with meniscus injuries and to explore its association with meniscus health. We hypothesize that vitamin D deficiency will be highly prevalent among patients with meniscus injuries and that low levels of 25-hydroxy vitamin D levels will be linked to poor meniscus health.

## 2. Materials and methods

### 2.1. Study Participants

This retrospective cohort study included patients hospitalized at Shanghai Tenth People’s Hospital with a diagnosis of meniscus injury between January 1 and December 31, 2023. In total, 198 meniscus injury patients were included in the study. The study included patients between 18 and 90 years old who had recently experienced knee pain or discomfort. Indicators of knee discomfort were confirmed through positive results on the McMurray’s test, Thessaly’s test or Joint line tenderness’s test. Participants with renal or liver failure, or any other chronic conditions, who were taking medications that might affect the metabolism of vitamin D, were not included in the study. Ethical approval was obtained from the ethics committee of Shanghai 10th People’s Hospital.

### 2.2. Clinical Data Collection

Clinical data was obtained from the electronic medical records including age, sex, blood pressure, body weight, glycated hemoglobin. The variety of vitamin data was also collected including vitamin B1, vitamin B2, niacin B3, pantothenic acid B5, vitamin B6, biotin B7, folate B9, vitamin B12, vitamin C, vitamin A, 25-hydroxy vitamin D, 25-hydroxy vitamin D2, 25-hydroxy vitamin D3, vitamin E and vitamin K1. The renal and hepatic functions were assessed by measuring ALT, AST, direct bilirubin, total bilirubin, total bile acid, urea, creatinine, uric acid, total cholesterol, triglyceride, high density lipoprotein and low-density lipoprotein. The numbers of inflammatory cells and levels of inflammatory markers were also collected. MRI scan analysis was based on the appearance of the meniscus on MRI images and was divided into four grades: Grade 0: Normal meniscus without any signs of tear or degeneration; Grade 1: Small, focus areas of increased signal within the meniscus on T2-weighted images, which do not extend to the surface; Grade2: Linear areas of increased signal within the meniscus on T2-weighted images, which also do not extend to the articular surface; Grade3: Linear areas of increased signal that extend to the articular surface of the meniscus on T2-weighted images.

### 2.3. Definition of vitamin D deficiency

Serum 25-hydroxy vitamin D is regarded as the most accurate indicator for evaluating vitamin D status (references). Given the varying optimal serum 25-hydroxy vitamin D levels for vitamin D deficiency adopted by different organizations, this study utilized the following classification: Vitamin D deficiency was defined as a 25-hydroxy vitamin D level of ≤30 nmol/L; Vitamin D sufficiency was defined as a 25-hydroxy vitamin D level of >30 nmol/L.

### 2.4. Statistical analysis

All data analyses were conducted using the GraphPad Prism 10. The normality of the data was assessed using the Kolmogorov-Smirnov test. For categorical variables, frequencies and percentages were reported, while continuous variables were summarized using mean and standard deviation or median and range. Continuous variables were assessed using unpaired t test with Welch’s correction or the Mann-Whitney U test depending on the data distribution. Spearman’s correlation coefficients were used to assess the correlations between 25-hydroxy vitamin D concentrations and parameters. Results were deemed statistically significant if the P-value was less than 0.05.

## 3. Results

### 3.1 Baseline Characteristics

The baseline characteristics of the study participants were outlined in Table 1. The cohort for the study comprised these 198 participants. At baseline, 162 patients (82%) had a 25(OH)D level < 30 nmol / L, while only 36 patients (18%) had a 25(OH)D level ≥ 30 nmol / L. When comparing characteristics between vitamin D deficiency group and vitamin D sufficiency group, there was no significant differences in age, body mass index, high blood pressure, low blood pressure, and glycated hemoglobin. Additionally, there was a significant difference in age between the female group, which was older than the male group (p = 0.0030).

**Table 1.**
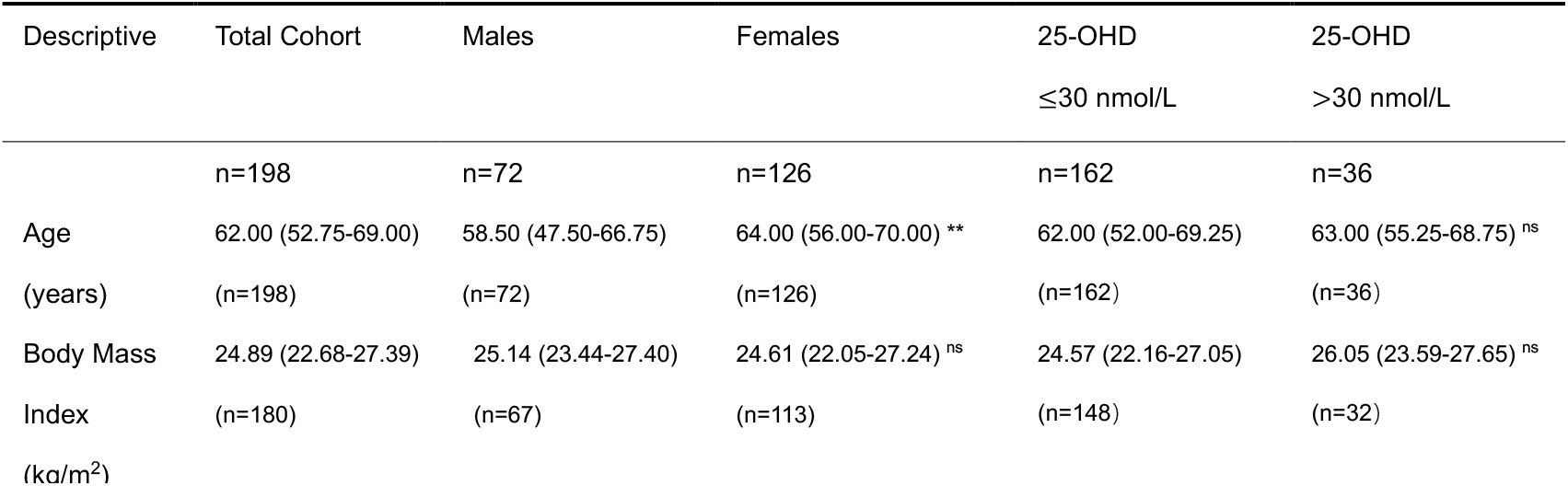
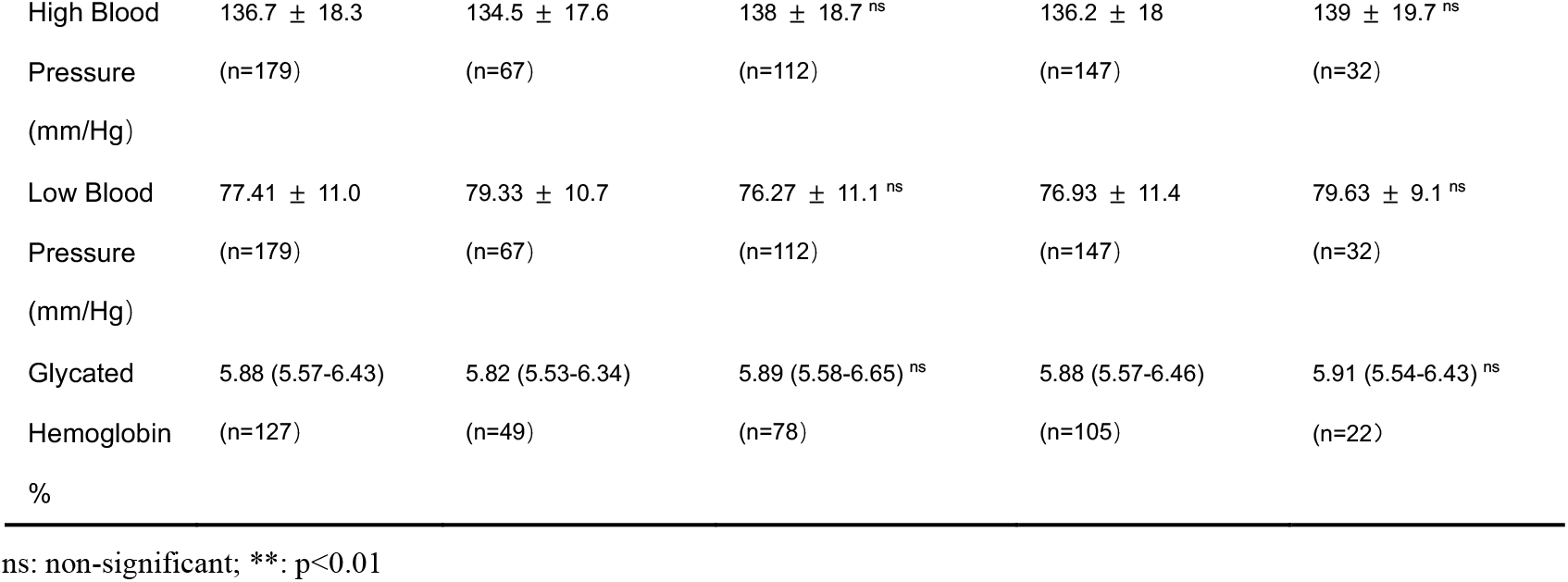
Baseline Characteristics of Meniscus Injury Patients.

### 3.2 Characteristics of Multivitamins in Groups with 25-hydroxy Vitamin D Deficiency and Sufficiency

We further investigated the levels of multiple vitamins in both the vitamin D deficiency and vitamin D sufficiency groups (Table 2). There were no significant differences in vitamin B1, vitamin B2, niacin B3, pantothenic acid B5, vitamin B6, biotin B7, folate B9, vitamin B12, vitamin C, vitamin A, 25-hydroxy vitamin D3, vitamin E, and vitamin K1 between the two groups. The mean levels of multiple vitamins were within the normal range based on clinical references in the participants, except for 25-hydroxy vitamin D, which shows 86% of patients are 25-hydroxy vitamin D deficient. This suggests that the level of 25-hydroxy vitamin D was more strongly associated with meniscus injury compared to other vitamins. Additionally, the levels of niacin B3, vitamin C and vitamin A were significantly lower in the female group compared to the male group (p=0.0189; p=0.0010; p=0.0119, respectively). The level of vitamin B1, folate B9 and vitamin E was significantly higher in the female group compared to the male group (p=0.0035; p=0.0001; p=0.0201, respectively). It is worth noting that the level of 25-hydroxy vitamin D2 was also higher in the 25-hydroxy vitamin D sufficient group compared to the deficient group (p=0.0225).

**Table 2.** Descriptive Vitamins Levels of Meniscus Injury Patients.

| Descriptive | Total Cohort | Males | Females | 25-OHD | 25-OHD |
| --- | --- | --- | --- | --- | --- |
|  |  |  |  | ≤30 nmol/L | >30 nmol/L |
| Vitamin B1 (ng/ml) | 2.02 (1.37-3.12)<br>(n=198) | 1.70 (0.96-2.76)<br>(n=72) | 2.19 (1.54-3.22) **<br>(n=126) | 2.00 (1.35-3.15)<br>(n=162) | 2.05 (1.42-2.95) <sup>ns</sup><br>(n=36) |
| Vitamin B2 (ng/ml) | 8.87 (5.93-13.51)<br>(n=198) | 8.56 (5.23-14.02)<br>(n=72) | 9.04 (5.97-13.30) <sup>ns</sup><br>(n=126) | 9.10 (6.00-13.51)<br>(n=162) | 8.70 (5.34-13.92) <sup>ns</sup><br>(n=36) |
| Niacin B3 (ng/ml) | 33.04 (16.55-57.20)<br>(n=198) | 38.71 (19.38-72.32)<br>(n=72) | 30.18 (15.18-49.43) *<br>(n=126) | 33.99 (16.93-58.50)<br>(n=162) | 28.09 (16.11-48.33) <sup>ns</sup><br>(n=36) |
| Pantothenic acid B5 (ng/ml) | 48.12 (39.01-61.16)<br>(n=198) | 48.36 (39.31-55.60)<br>(n=72) | 47.88 (38.23-64.23) <sup>ns</sup><br>(n=126) | 48.12 (38.62-59.82)<br>(n=162) | 48.57 (43.00-66.54) <sup>ns</sup><br>(n=36) |
| Vitamin B6<br>(ng/ml) | 2.78 (2.01-8.00)<br>(n=198) | 2.57 (1.82-6.87)<br>(n=72) | 3.09 (2.16-8.45) <sup>ns</sup><br>(n=126) | 2.75 (2.01-8.00)<br>(n=162) | 3.91 (2.09-8.01) <sup>ns</sup><br>(n=36) |
| Biotin B7<br>(ng/ml) | 0.30 (0.22-0.49)<br>(n=198) | 0.29 (0.22-0.46)<br>(n=72) | 0.31 (0.22-0.51) <sup>ns</sup><br>(n=126) | 0.30 (0.21-0.50)<br>(n=162) | 0.28 (0.22-0.46) <sup>ns</sup><br>(n=36) |
| Folate B9<br>(ng/ml) | 12.93 (8.76-21.12)<br>(n=198) | 11.14 (6.92-16.38)<br>(n=72) | 15.50 (10.08-25.63) ***<br>(n=126) | 12.93 (9.07-21.48)<br>(n=162) | 13.72 (7.22-20.45) <sup>ns</sup><br>(n=36) |
| Vitamin B12<br>(ng/ml) | 0.52 (0.41-0.70)<br>(n=198) | 0.50 (0.42-0.67)<br>(n=72) | 0.53 (0.41-0.78) <sup>ns</sup><br>(n=126) | 0.52 (0.42-0.68)<br>(n=162) | 0.53 (0.39-0.81) <sup>ns</sup><br>(n=36) |
| Vitamin C<br>(µg / ml) | 8.06 (6.49-10.43)<br>(n=198) | 9.09 (7.13-10.88)<br>(n=72) | 7.70 (6.30-9.39) **<br>(n=126) | 8.06 (6.47-10.51)<br>(n=162) | 8.05 (6.51-9.60) <sup>ns</sup><br>(n=36) |
| Vitamin A<br>(ng / ml) | 476.5 (393.9-583.5)<br>(n=198) | 502.7 (418.4-645.9)<br>(n=72) | 465.6 (383.6-554.6) *<br>(n=126) | 467.4 (390.2-578.4)<br>(n=162) | 504.3 (404.7-613.4) <sup>ns</sup><br>(n=36) |
| 25-Hydroxy<br>Vitamin D<br>(ng / ml) | 21.90 (17.16-28.09)<br>(n=198) | 21.69 (16.11-26.32)<br>(n=72) | 22.77 (17.24-28.93) <sup>ns</sup><br>(n=126) | 20.58 (15.56-24.18)<br>(n=162) | 36.46 (32.12-39.36) ****<br>(n=36) |
| 25-Hydroxy<br>Vitamin D2<br>(ng / ml) | 1.11 (0.76-1.54)<br>(n=198) | 1.08 (0.77-1.43)<br>(n=72) | 1.14 (0.74-1.70) <sup>ns</sup><br>(n=126) | 1.07 (0.75-1.48)<br>(n=162) | 1.38 (0.91-2.21) *<br>(n=36) |
| 25-Hydroxy<br>Vitamin D3<br>(ng / ml) | 20.63 (15.07-26.58)<br>(n=198) | 21.00 (16.12-26.98)<br>(n=72) | 20.11 (13.97-26.20) <sup>ns</sup><br>(n=126) | 20.27 (15.46 -26.20)<br>(n=162) | 20.77 (14.13-27.05) <sup>ns</sup><br>(n=36) |
| Vitamin E<br>(µg / ml) | 11.30 (9.20-13.66)<br>(n=198) | 10.50 (8.64-12.85)<br>(n=72) | 11.63 (9.64-13.86) *<br>(n=126) | 11.16 (9.12-13.51)<br>(n=162) | 11.77 (9.69-14.37) <sup>ns</sup><br>(n=36) |
| Vitamin K1<br>(ng / ml) | 1.32 (0.69-2.47)<br>(n=198) | 1.41 (0.69-2.85)<br>(n=72) | 1.23 (0.69-2.21) <sup>ns</sup><br>(n=126) | 1.30 (0.71-2.42)<br>(n=162) | 1.50 (0.42-2.82) <sup>ns</sup><br>(n=36) |

ns: non-significant; \*: p<0.05; \*\*: p<0.01; \*\*\*\*: p<0.0001

### 3.3 Characteristics of Biochemical Analysis in Groups with 25-hydroxy Vitamin D Deficiency and Sufficiency

Table 3 shows that there were no significant differences in ALT, AST, direct bilirubin, total bilirubin, total bile acid, urea, creatinine, uric acid, total cholesterol, triglyceride, high-density lipoprotein, and low-density lipoprotein between the 25-hydroxy vitamin D deficiency and sufficiency groups. There was also no significant difference regarding electrolytes between the two groups. The levels of ALT, total bilirubin, creatinine, uric acid and K were significantly lower in females group compared to the male group (p=0.0038; p=0.0313; p<0.0001; p<0.0001, respectively). The levels of P, Total cholesterol and high-density lipoprotein concentrations were significantly higher in females compared with males (p=0.0013; p=0.0004; p<0.0001, respectively).

**Table 3.** Clinical Results of Meniscus Injury Patients.

| Descriptive | Total Cohort | Males | Females | 25-OHD | 25-OHD |
| --- | --- | --- | --- | --- | --- |

|  |  |  |  | ≤30 nmol/L | > 30 nmol/L |
| --- | --- | --- | --- | --- | --- |
| ALT (U/L) | 20.70 (14.15-29.70)<br>(n=158) | 23.45 (18.78-31.45)<br>(n=54) | 18.15 (13.20-26.80) **<br>(n=104) | 20.60 (13.90-29.38)<br>(n=128) | 22.05 (15.18-31.38) <sup>ns</sup><br>(n=30) |
| AST (U/L) | 19.20 (15.70-23.55)<br>(n=157) | 19.75 (16.28-24.13)<br>(n=54) | 18.60 (15.70-23.30) <sup>ns</sup><br>(n=103) | 19.40 (16.00-23.80)<br>(n=127) | 18.40 (14.05-23.88) <sup>ns</sup><br>(n=30) |
| Direct Bilirubin<br>(μmol /L) | 3.60 (2.80-4.93)<br>(n=146) | 3.75 (3.00-5.18)<br>(n=52) | 3.40 (2.70-4.80) <sup>ns</sup><br>(n=94) | 3.60 (2.80-5.00)<br>(n=118) | 3.65 (2.65-4.60) <sup>ns</sup><br>(n=28) |
| total bilirubin<br>(μmol / L) | 11.60 (8.48-16.35)<br>(n=144) | 12.95 (9.80-17.73)<br>(n=52) | 10.80 (8.00-16.00) *<br>(n=92) | 11.60 (8.70-16.55)<br>(n=117) | 12.40 (7.80-14.70) <sup>ns</sup><br>(n=27) |
| total bile acid<br>(μmol /L) | 4.10 (2.75-7.25)<br>(n=108) | 4.10 (2.90-8.40)<br>(n=39) | 4.10 (2.65-7.10) <sup>ns</sup><br>(n=69) | 4.05 (2.85-7.63)<br>(n=86) | 4.65 (2.68-6.30) <sup>ns</sup><br>(n=22) |
| Urea<br>(mmol / L) | 5.60 (4.55-6.40)<br>(n=157) | 5.60 (4.50-6.65)<br>(n=53) | 5.35 (4.53-6.28) <sup>ns</sup><br>(n=104) | 5.40 (4.30-6.50)<br>(n=127) | 5.65 (4.90-5.93) <sup>ns</sup><br>(n=30) |
| Creatinine<br>(μmol / L) | 61.00 (51.00-72.00)<br>(n=158) | 75.00 (67.75-79.25)<br>(n=54) | 54.50 (48.25-61.75) ****<br>(n=104) | 63.00 (51.25-72.75)<br>(n=128) | 56.00 (48.75-68.25) <sup>ns</sup><br>(n=30) |
| Uric Acid<br>(μmol / L) | 329.0 (276.5-387.5)<br>(n=157) | 373.5 (328.8-452.0)<br>(n=54) | 307.0 (258.0-348.0) ****<br>(n=103) | 329.0 (274.0-388.0)<br>(n=127) | 329.5 (277.8-387.3) <sup>ns</sup><br>(n=30) |
| K (mmol / L) | 4.13 (3.96-4.29)<br>(n=158) | 4.23 (4.01-4.44)<br>(n=54) | 4.10 (3.92-4.22) ***<br>(n=104) | 4.14 (3.95-4.34)<br>(n=128) | 4.13 (3.95-4.19) <sup>ns</sup><br>(n=30) |
| Na (mmol / L) | 143.0 (142.0-144.0)<br>(n=158) | 142.5 (142.0-144.0)<br>(n=54) | 143.0 (142.0-144.0) <sup>ns</sup><br>(n=104) | 143.0 (142.0-144.0)<br>(n=128) | 143.0 (141.0-144.0) <sup>ns</sup><br>(n=30) |
| Cl (mmol / L) | 109.0 (107.0-110.0)<br>(n=158) | 109.0 (106.8-110.0)<br>(n=54) | 109.0 (107.0-111.0) <sup>ns</sup><br>(n=104) | 109.0 (107.0-110.8)<br>(n=128) | 109.0 (107.0-110.0) <sup>ns</sup><br>(n=30) |
| Ca (mmol / L) | 2.35 (2.29-2.41)<br>(n=158) | 2.36 (2.30-2.41)<br>(n=54) | 2.34 (2.29-2.41) <sup>ns</sup><br>(n=104) | 2.35 (2.29-2.41)<br>(n=128) | 2.36 (2.30-2.41) <sup>ns</sup><br>(n=30) |
| P (mmol / L) | 1.12±0.2<br>(n=158) | 1.05±0.2<br>(n=54) | 1.15±0.2**<br>(n=104) | 1.12±0.2<br>(n=128) | 1.12±0.2<sup>ns</sup><br>(n=30) |
| Mg (mmol / L) | 0.87±0.1<br>(n=158) | 0.87±0.1<br>(n=54) | 0.86±0.1<sup>ns</sup><br>(n=104) | 0.87±0.1<br>(n=128) | 0.86±0.1<sup>ns</sup><br>(n=30) |
| Total Cholesterol<br>(mmol / L) | 5.13±1.0<br>(n=88) | 4.64±0.9<br>(n=33) | 5.43±1.0***<br>(n=55) | 5.12±1.0<br>(n=71) | 5.20±1.0<sup>ns</sup><br>(n=17) |
| Triglyceride (mmol / L) | 1.70 (1.13-2.35)<br>(n=88) | 1.76 (1.15-2.57)<br>(n=33) | 1.64 (1.10-2.33) <sup>ns</sup><br>(n=55) | 1.62 (0.97-2.37)<br>(n=71) | 2.04 (1.38-2.18) <sup>ns</sup><br>(n=17) |
| High Density<br>Lipoprotein (mmol / L) | 1.21 (1.04-1.39)<br>(n=87) | 1.05 (0.96-1.24)<br>(n=32) | 1.30 (1.11-1.47) ****<br>(n=55) | 1.21 (1.04-1.34)<br>(n=70) | 1.23 (1.11-1.41) <sup>ns</sup><br>(n=17) |
| Low Density Lipoprotein<br>(mmol / L) | 3.18±0.9<br>(n=87) | 2.96±0.8<br>(n=32) | 3.31±0.9 <sup>ns</sup><br>(n=55) | 3.16±0.8<br>(n=70) | 3.27±1.0 <sup>ns</sup><br>(n=17) |

ns: non-significant; \*: p<0.05; \*\*: p<0.01; \*\*\*: p<0.001; \*\*\*\*: p<0.0001; ALT: Alanine aminotransferase; AST: Aspartate aminotransferase.

### 3.4 Characteristics of Immune Cells in Groups with 25-hydroxy Vitamin D Deficiency and Sufficiency

Table 4 demonstrates that there were no significant differences between the 25-hydroxy vitamin D deficiency and sufficiency groups in terms of CRP, WBCs, neutrophils, lymphocytes, monocytes, eosinophils, and basophils. Moreover, females had significantly lower levels of WBCs, neutrophils, monocytes, eosinophils and basophils compared to males (p=0.0071; p=0.0305; p=0.0276; p=0.0140; p=0.0267, respectively).

**Table 4.** Clinical Results of Inflammatory Cells Numbers of Meniscus Injury Patients.

| Descriptive | Total Cohort | Males | Females | 25-OHD | 25-OHD |
| --- | --- | --- | --- | --- | --- |
| | | | | $\leq 30$ nmol/L | $> 30$ nmol/L |
| CRP (mg / L) | 1.46 (0.70-4.35) | 1.44 (0.67-5.10) | 1.47 (0.73-4.06) <sup>ns</sup> | 1.46 (0.70-3.89) | 1.48 (0.62-5.42) <sup>ns</sup> |
|  | (n=115) | (n=42) | (n=73) | (n=99) | (n=16) |
| WBC | 6.42 (5.28-7.67) | 6.95 (5.72-8.23) | 6.07 (5.11-7.49) ** | 6.40 (5.25-7.55) | 6.55 (5.43-8.35) <sup>ns</sup> |
| ($10^9$ / L) | (n=197) | (n=72) | (n=125) | (n=162) | (n=35) |
| Neutrophil | 3.93 (3.13-5.07) | 4.20 (3.34-5.46) | 3.76 (2.96-4.87) * | 3.87 (3.11-5.05) | 4.19 (3.13-5.39) <sup>ns</sup> |
| ($10^9$ / L) | (n=197) | (n=72) | (n=125) | (n=162) | (n=35) |
| Lymphocyte | 1.74 (1.41-2.08) | 1.76 (1.38-2.24) | 1.70 (1.43-2.04) <sup>ns</sup> | 1.74 (1.41-2.06) | 1.78 (1.43-2.26) <sup>ns</sup> |
| ($10^9$ / L) | (n=197) | (n=72) | (n=125) | (n=162) | (n=35) |
| Monocyte | 0.38 (0.29-0.52) | 0.43 (0.30-0.57) | 0.36 (0.27-0.48) * | 0.38 (0.28-0.51) | 0.36 (0.29-0.55) <sup>ns</sup> |
| ($10^9$ / L) | (n=197) | (n=72) | (n=125) | (n=162) | (n=35) |
| Eosinophil | 0.09 (0.05-0.15) | 0.11 (0.06-0.21) | 0.08 (0.04-0.13) * | 0.09 (0.05-0.14) | 0.09 (0.05-0.20) <sup>ns</sup> |
| ($10^9$ / L) | (n=186) | (n=66) | (n=120) | (n=153) | (n=33) |
| Basophil | 0.03 (0.02-0.04) | 0.03 (0.02-0.04) | 0.03 (0.02-0.04) * | 0.03 (0.02-0.04) | 0.03 (0.02-0.04) <sup>ns</sup> |
| ($10^9$ / L) | (n=195) | (n=71) | (n=124) | (n=160) | (n=35) |

ns: non-significant; \*: $p<0.05$ ; \*\*: $p<0.01$ ; CRP: C-reactive protein; WBC: White blood cells.

### 3.5 Correlation between 25-Hydroxy Vitamin D Concentration and Parameters

Table 5 indicates that there was no significant correlation between 25-hydroxy vitamin D Concentration and age, BMI, high blood pressure, low blood pressure, glycated hemoglobin, total cholesterol, triglyceride, high-density lipoprotein, low-density lipoprotein, and CRP.

**Table 5.**
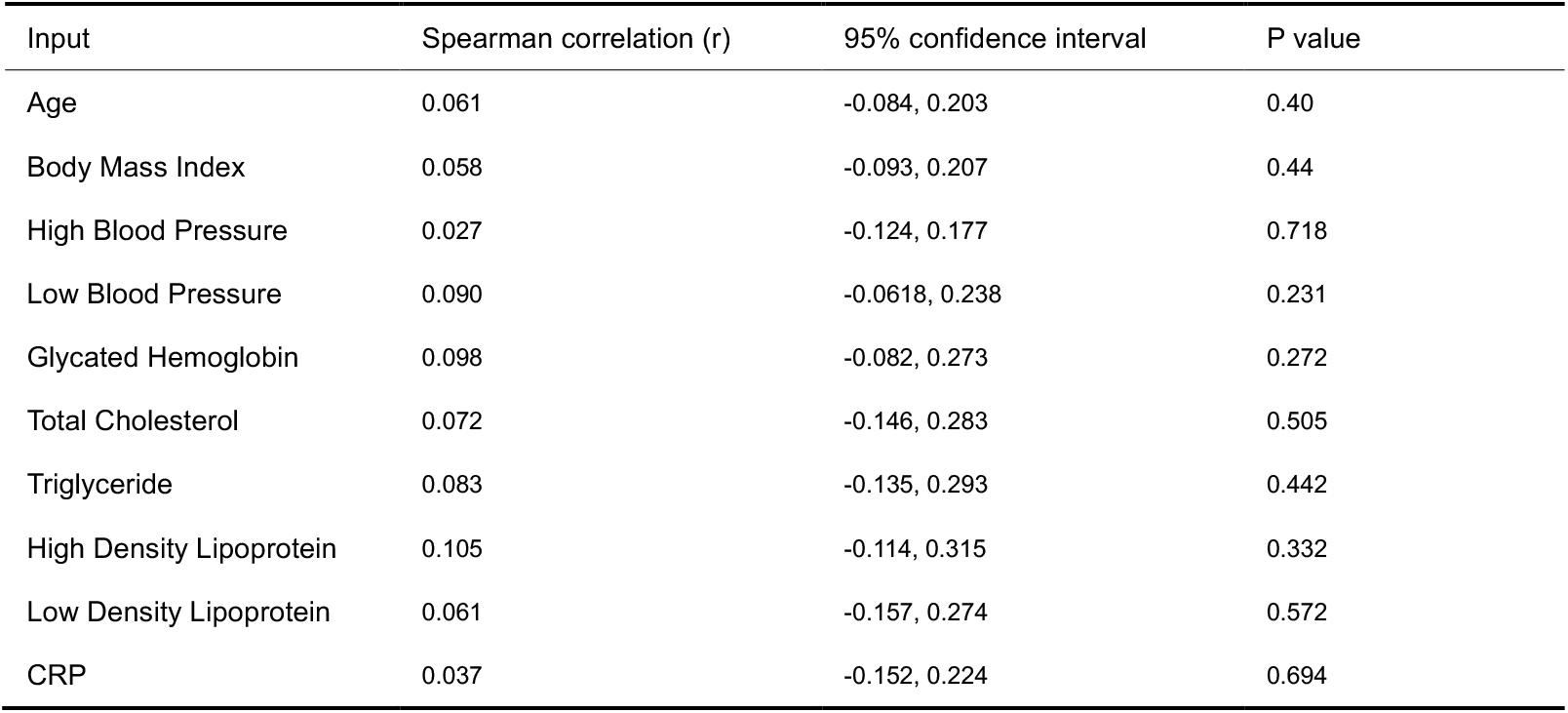
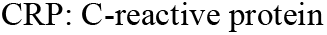
Correlation between Clinical Characteristics and 25-hydroxy Vitamin D Status.

| Input | Spearman correlation (r) | 95% confidence interval | P value |
| --- | --- | --- | --- |
| Age | 0.061 | -0.084, 0.203 | 0.40 |
| Body Mass Index | 0.058 | -0.093, 0.207 | 0.44 |
| High Blood Pressure | 0.027 | -0.124, 0.177 | 0.718 |
| Low Blood Pressure | 0.090 | -0.0618, 0.238 | 0.231 |
| Glycated Hemoglobin | 0.098 | -0.082, 0.273 | 0.272 |
| Total Cholesterol | 0.072 | -0.146, 0.283 | 0.505 |
| Triglyceride | 0.083 | -0.135, 0.293 | 0.442 |
| High Density Lipoprotein | 0.105 | -0.114, 0.315 | 0.332 |
| Low Density Lipoprotein | 0.061 | -0.157, 0.274 | 0.572 |
| CRP | 0.037 | -0.152, 0.224 | 0.694 |

CRP: C-reactive protein

### 3.6 Patients with Severe Meniscus Injury Present a 25-Hydroxy Vitamin D Deficiency

According to the MRI report, out of 198 patients, the MRI reports of 127 patients met the criteria for this study. These MRI reports were divided into two groups based on meniscus injury classifications: (i) patients with meniscal injuries greater than or equal to grade 2; and (ii) patients with meniscal injuries less than grade 2. Our results show that the level of 25-hydroxy vitamin D was lower in the group with meniscal injury greater than or equal to grade 2 group compared to the group with meniscal injuries less than grade 2 (Figure 1).

**Figure 1.**
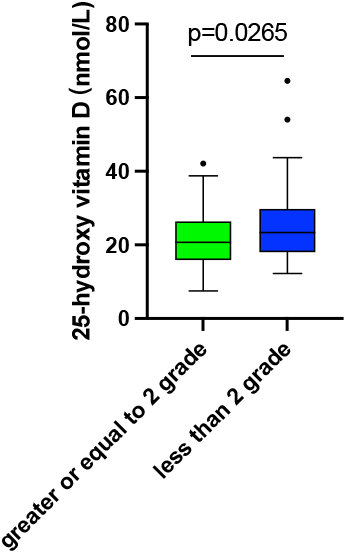
Comparison of 25-hydroxy vitamin D levels of between both groups by stratified MRI results

## 4. Discussion

This study is the first to demonstrate a significant prevalence of 25-hydroxy vitamin D deficiency among patients with meniscus injuries. Furthermore, we observed that patients with more severe meniscus injuries have lower 25-hydroxy vitamin D levels. It is worth noting that in our study, we did not observe any correlation between 25-hydroxy vitamin D levels and baseline characters or inflammatory biomarkers. It suggests that patients with meniscus injury may affect the absorption and metabolism of 25-hydroxy vitamin D, independent of age, gender, BMI, and inflammatory response. As in our study, the observation that lower 25-hydroxy vitamin D concentrations were associated with more severe meniscus injuries was consistently supported by our MRI results. To our knowledge, none of the observation studies conducted so far have reported evidence of a relationship between 25-hydroxy vitamin D and meniscus injuries.

Several potential mechanisms could explain the protective effect of higher 25-hydroxy vitamin D levels on meniscus health. Vitamin D is essential for maintaining bone health and contributes to muscle function. Several studies have shown that vitamin D promotes calcium absorption and helps maintain bone density [8-9], which could reduce the risk of joint injuries, including meniscal tears. Furthermore, growing evidence indicates that vitamin D could be crucial for muscle damage repair and regeneration [16-17]. Mechanistically, muscle cells express the vitamin D receptor, as is CYP27B1, which converts 25(OH)D into its active form, 1,25(OH)D. *In vitro* and *in vivo* rodent studies indicate that vitamin D reduces the production of reactive oxygen species, boosts antioxidant capacity, and prevents oxidative stress, thereby preventing muscle damage [18]. Recently, a mendelian randomization study has shown that a causal relationship between vitamin D and skeletal muscle health [19]. Effective strategies for preventing vitamin D deficiency may help mitigate muscle weakness, thereby improving joint stability and reducing the risk of meniscus injury.

Knee pain is a typical feature of meniscus injury patients. Most patients with a meniscus injury seek medical attention due to knee pain. While there is controversy surrounding the relationship between vitamin D levels and knee pain. Some studies suggest that adequative vitamin D levels can help alleviate knee pain and improve joint health [20-21], while others find no significant correlation [22-23]. Based on our cohort study, we observed that patients suffering from chronic knee pain have lower 25-hydroxy vitamin D levels (<30 nmol/L), this suggests that the concentration of vitamin D may be related to the duration of knee pain in patients with meniscus injuries. Recently, a cross-section study of 349,221 adults in the UK has shown that severe vitamin D deficiency (<25nmol/L) is independently associated with chronic widespread pain [24]. Therefore, vitamin D assessment and treatment may need be further exploration in patients suffering from pain.

Our study has several strengths, including a well-characterized cohort and the use of MRI to accurately assess the severity of meniscus injury. However, it is important to acknowledge several limitations in this study. Firstly, serum 25-hydroxy vitamin D concentrations were measured only once at baseline, preventing us from tracking the long-term trajectories of serum 25-hydroxy vitamin D concentrations during the follow-up periods. Secondly, this study did not evaluate the effects of vitamin D supplements on meniscus injuries. Thirdly, the small sample size may limit the power to detect significant associations. Fourthly, this study did not adjust potential confounding factors, such as diet, sun exposure, and physical activity, which may influence the results.

## 5. Conclusions

In conclusion, the observation study is the first to highlight the potential relationship between serum25-hydroxy vitamin D levels and meniscus injury. While our findings suggest that 25-hydroxy vitamin D deficiency is associated with meniscus injury, further research is needed to establish causality and explore the underlying mechanisms.

## Data Availability

All data produced in the present study are available upon reasonable request to the authors

## Author Contributions

Conceptualization, S.H. and C.J; data collection, ZZ; formal analysis, S.H; writing-original draft preparation, S.H; writing-review and editing, S.H, ZZ, and C.J; All authors have read and agreed to the published version of the manuscript.

